# “Sleepless in Lockdown”: unpacking differences in sleep loss during the coronavirus pandemic in the UK

**DOI:** 10.1101/2020.07.19.20157255

**Authors:** Jane Falkingham, Maria Evandrou, Min Qin, Athina Vlachantoni

**Affiliations:** ESRC Centre for Population Change, University of Southampton, Southampton, UK; Centre for Research on Ageing, University of Southampton, Southampton, UK

**Keywords:** COVID-19, Pandemic, Sleep Loss, Mental Health, Gender, Ethnicity

## Abstract

**Background:** Covid-19 has been shown to be having a disproportionate impact on the health of individuals from different ethnic groups and those employed in certain occupations, whilst the indirect impacts of Covid-19, including the closure of schools and business and the move to home working, fall disproportionately on the young and on women. These factors may in turn impact upon sleep health. Research on sleep deprivation during the pandemic crisis to date has been limited. The present study aimed to explore the levels and social determinants of self-reported sleep loss among the general population during the Covid-19 pandemic in the UK, with a particular focus on ethnic and gender disparities.

**Methods:** Newly available national representative survey data from Understanding Society COVID- 19 Study collected during April 2020 were analysed. These data were linked to Wave 9 of Understanding Society conducted in 2018/19, providing information about the respondents prior to the outbreak of the pandemic. Cross-sectional analysis provided prevalence estimates, whilst analysis of the linked longitudinal data provided incidence estimates. The analytical sample included 15,360 respondents aged 16 and above; among these, 12,206 reported no problem of sleep loss before the epidemic.

**Results:** Prevalence and incidence rates of perceived sleep loss were 24.7% and 20.2% respectively. Women (at the level of 31.8% and 27.0%) and individuals from Black, Asian, and minority ethnic (BAME) communities (at the level of 32.0% and 24.6%) were more vulnerable to sleep deprivation due to the pandemic. Multivariate regression analysis shows that being female, the presence of young children in the household, perceived financial difficulties and being a Covid-19-related key worker were all predictive of sleep loss. Once these covariates were controlled for the bivariate relationship between ethnicity and sleep loss was reversed, reflecting the complex interaction between the coronavirus epidemic and ethnicity.

**Conclusions:** The pandemic has widened the disparity of sleep deprivation across different groups, with women with young children, key workers and people of BAME heritage all experiencing difficulty in sleeping, which in turn may negatively affect mental and physical health and well-being.

## Introduction

The coronavirus disease (Covid-19) is impacting upon physical and mental health globally. Sleep problems associated with increased psychosocial stressors induced by the coronavirus itself, and as a result of the social distancing measures that have been introduced to manage the virus, are emerging as a significant outcome of the Covid-19 crisis. According to a recent report, more than half of the UK population has struggled with sleep during the lockdown (1). Sleep has long been recognised as an essential determinant of human health and performance. Good sleep restores energy, promotes healing, interacts with the immune system and impacts upon behaviour (2). Even acute sleep deprivation can impair judgement and cognitive performance, while persistent deviations have been linked to disease development and increased mortality (3, 4). During the pandemic, lack of sleep may itself have had knock-on effects on people’s capacity to be resilient. However, to date relatively limited research has been conducted on sleep deprivation during the pandemic.

Sleep is known to be regulated by circadian rhythms, sleep-wake homoeostasis and cognitive-behavioural influences (3). With regards to environmental, behavioural and health determinants, poor sleep has been associated with stress, anxiety, work pressures, financial concerns, mental and physical impairments, and physical activity (5, 6, 7, 8). Previous studies have found that women were more likely than men to have trouble falling and staying asleep frequently, or to have insufficient sleep (9, 10). The relationship between ethnicity and sleep is complicated due to the broader social and environmental factors determining group differences in sleep behaviours and the structural relationships between these factors and ethnicity (11). Some studies reported that inadequate sleep duration and poorer sleep were more prevalent among low-income and black and minority ethnic (BAME) communities (12), whereas others have failed to find this association (13).

In understanding the relationship between Covid-19 and sleep, it is helpful to conceptually distinguish between those factors linked to being *infected* with Covid-19 and those associated with the policy responses and measures introduced to manage the pandemic that have *affected* everyday life. Although it is still relatively early in our understanding of Covid-19, research by Public Health England 2020 has highlighted that older people, men and individuals from BAME groups are all at increased risk of developing a severe response to the virus and to die from it (14). The reasons for the heightened risk amongst certain ethnic groups remains unresolved, but potential contributors include the disproportionate representation of BAME individuals in some high risk occupations including front line health care work, as well as wider environmental factors that, interwoven with issues of inequality, deprivation and structural racism, manifest in longstanding ethnic disparities in health (15). A priori, we might expect those groups facing the greatest health risks from the virus to report increased sleep loss due to worry and thus to observe differences across ethnic groups.

The public health actions taken to control the spread of the virus have, however, impacted all domains of life and thus affected all individuals. On 23rd March 2020, the UK went into lockdown in an unprecedented attempt to limit the spread of coronavirus, with the Government mandating all those who could to work at home, closing schools, restaurants and all but essential shops, and advising the population to stay at home and limit contact with other individuals outside their household. The resultant move to home working and learning and, for some, the loss of work altogether, along with limited social contact and increased isolation, may all be anticipated to affect mental well-being and the ability to sleep. Preliminary evidence points towards the young and women being disproportionally affected, with women being more likely than men to be working in sectors that were locked down (16) and mothers being more likely to be interrupted whilst working from home than fathers (17). Lockdown has also resulted in increased instances of domestic violence; the UK domestic abuse organisation, Refuge, reported a 25 per cent increase in calls and online requests since the lockdown began in March 2020 (18). Given this, we might anticipate a gender differential in increased sleep loss, with women being disproportionately affected by lockdown compared with men.

This study aims to provide novel evidence regarding patterns of self-reported increased sleep loss due to worry during the first four weeks of the Covid-19 related lockdown in the UK. Using recently collected nationally representative survey data, the research provides the first estimates of the prevalence and incidence of increased sleep loss since the coronavirus pandemic. It attempts to unpack the impact of factors associated with being *infected* and being *affected* by Covid-19, with a particular focus on the extent to which the pandemic has exacerbated differentials in sleep loss by ethnicity and gender.

## Data and methods

### Data and study population

This study used data drawn from the first wave of Understanding Society Covid-19 Study, conducted in April 2020 (19). Fieldwork was completed on 29^th^ April and thus covers the first month of lockdown in the UK. The data were linked to Wave 9 of Understanding Society conducted in 2018/19 (20), providing information about the respondents prior to the outbreak of the pandemic. Cross-sectional analysis of the Covid-19 Study was used for prevalence estimates and longitudinal analysis of the linked data was used for incidence estimates. The analytical sample was all respondents aged 16 and over, constituting a sample size of 15,360; among them, 12,206 reported no sleep loss before the epidemic. The characteristics of the sample are shown in **Table 1**.

**Table 1.** Sample characteristics.

|  | % | Number of respondents |
| --- | --- | --- |
| **Total** | **100.0** | **15360** |
| **Age group** |  |  |
| 16-24 | 8.5 | 895 |
| 25-44 | 26.9 | 3966 |
| 45-64 | 38.8 | 6493 |
| 65-74 | 16.8 | 2823 |
| 75+ | 9.0 | 1183 |
| **Sex** |  |  |
| Men | 46.1 | 6406 |
| Women | 53.9 | 8954 |
| **Ethnicity** |  |  |
| British/English/Scottish/Welsh/Northern Irish (white) | 88.8 | 13023 |
| Other white | 4.2 | 708 |
| Black, Asian, and minority ethnic (BAME) | 7.0 | 1629 |
| **Highest qualification** |  |  |
| No qualification | 15.8 | 2015 |
| GCSE or lower | 25.8 | 3170 |
| A level | 11.7 | 1495 |
| Degree | 37.0 | 6115 |
| Unknown | 9.7 | 2565 |
| **Live with a partner** |  |  |
| No | 32.8 | 4175 |
| Yes | 67.2 | 11185 |
| **Children in the house** |  |  |
| At least one child aged 0-4 | 8.4 | 1418 |
| At least one school children aged 5-18 | 18.5 | 3036 |
| Others | 73.1 | 10906 |
| **Key worker** |  |  |
| No | 33.7 | 5199 |
| Yes | 26.9 | 4216 |
| Not in paid or self-employed work | 39.4 | 5945 |
| **Has had symptoms that could be coronavirus** |  |  |
| No | 88.2 | 13456 |
| Yes | 11.8 | 1904 |
| **Feel lonely** |  |  |
| Hardly ever | 62.2 | 9870 |
| Sometime | 29.3 | 4383 |
| Often | 8.5 | 1107 |
| **Subjective current financial situation** |  |  |
| Living comfortably | 33.1 | 5497 |
| Doing alright | 43.0 | 6610 |
| Just about getting by | 17.8 | 2427 |
| Finding it quite difficult/very difficult | 6.1 | 826 |
| **Subjective future financial situation** |  |  |
| Better off | 7.7 | 1131 |
| Worse off | 16.8 | 2620 |
| About the same | 75.5 | 11609 |
| **Smoker** |  |  |
| No | 89.3 | 14029 |
| Yes | 10.7 | 1331 |
| **Physical activity** |  |  |
| No exercise | 9.9 | 1349 |
| Had some exercise but not meet NHS guidance | 40.3 | 6215 |
| Had exercise meet NHS guidance | 49.8 | 7796 |
| **Prior problem of sleep** |  |  |
| No | 83.1 | 12206 |
| Yes | 15.7 | 2183 |
| Unknown | 1.3 | 971 |
| **Region** |  |  |
| North East | 4.1 | 529 |
| North West | 11.1 | 1495 |
| Yorkshire and The Humber | 8.6 | 1302 |
| East Midlands | 7.9 | 1178 |
| West Midlands | 8.9 | 1293 |
| East of England | 10.3 | 1492 |
| London | 10.2 | 1517 |
| South East | 14.7 | 2178 |
| South West | 9.5 | 1461 |
| Wales | 4.6 | 916 |
| Scotland | 7.9 | 1358 |
| Northern Ireland | 2.2 | 641 |

Source: authors' analysis, Understanding Society: COVID-19 Study, 2020.
Note: All proportions are weighted using sample weights. Number of respondents are unweighted.

### Dependent variables

**The outcome variables** included (i) whether the respondent reported an increase in sleep loss over worry in the last few weeks and (ii) a new occurrence of the problem of sleep loss as compared with the situation prior to the coronavirus pandemic.

The question on sleep loss was identical across both the Understanding Society COVID-19 Study and Wave 9 of Understanding Society. The specific question wording, along with the four response categories, is presented in the text box below.

*The next questions are about how you have been feeling over the last few weeks*.

*Have you recently lost much sleep over worry?*

1. *Not at all*
2. *No more than usual*
3. *Rather more than usual*
4. *Much more than usual*

Source: University of Essex (2020).

For the purposes of the main analysis we group the first two options as indicating no problem of increased sleep loss due to worry and the latter two options as indicating a problem of sleep loss. A respondent was defined as experiencing sleep loss during the lockdown if he or she reported sleep loss over worry in the last few weeks ‘rather more than usual’ or ‘much more than usual’ in the Understanding Society COVID-19 Study. A respondent was defined as experiencing a new occurrence of sleep loss due to worry if he or she was *not* classified as having a problem of sleep loss at Wave 9 of Understanding Society but subsequently reported sleep loss in the Understanding Society COVID-19 Study. Both outcome variables were binary (1=yes; 0=no).

### Independent variables

A range of explanatory variables were included in the analyses, reflecting both known associates of sleep loss (9, 21) as well as those that we hypothesis may be associated with heightened anxiety during the pandemic. Demographic and socioeconomic characteristics included age, gender, ethnicity and educational qualification. Gender distinguished between men and women; the survey responses do not differentiate those whose reported sex has changed since birth or those who classify themselves as intersex. Ethnicity was classified into three groups: British/English/Scottish/Welsh/Northern Irish (white), Other white, and Black, Asian, and minority ethnic (BAME).

Health risk behaviours included whether the respondent was a current smoker and recent physical activities^1^ classified into three categories (no exercise, had some exercise but did not meet NHS guidance, and had exercise meeting or exceeding NHS guidance).

Variables capturing factors associated with Covid-19 itself included whether the respondent reported having experienced symptoms that could be coronavirus and being a key worker^2^. Other variables aimed to capture the impacts of the policy response to Covid-19, particularly the effect of lockdown. Increased stress related to childcare and home schooling was proxied by the presence of children in the house (whether at least one child aged 0-4 or school-aged child), and whether the respondent was living with a partner. Exposure to financial stress was proxied by two variables capturing the respondents subjective view of their current and future financial situation. Social isolation was measured by frequency of feeling lonely.

Finally, in order to capture regional differences in the intensity of the pandemic^3^, a variable reflecting the respondents’ place of respondence was included, based on the UK government office region (North East, North West, Yorkshire and The Humber, East Midlands, West Midlands, East of England, London, South East, South West, Wales, Scotland, and Northern Ireland).

### Analytical approach

Regression analysis, among the total population and then for men and women separately, was used to assess the existence and strength of associations between sleep loss as well as the new occurrence of sleep loss, and coronavirus related circumstances during the pandemic.

## Results

### Bi-variate analysis: prevalence and incidence rate of perceived sleep loss during the coronavirus pandemic

One in four people in the UK reported increased sleep loss due to worry during the first four weeks of the coronavirus pandemic lockdown in the UK (**Table 2**). There were clear differences between women and men (Figure 1), by age group (Figure 2) and across ethnic communities (Figure 3). Strikingly, although men have been found to face a higher risk of experiencing severe symptoms and dying from Covid-19, women were nearly twice as likely as men to report that they had lost sleep ‘much more than usual’ (6.2% v 3.2%) and ‘rather more than usual’ (25.6% v 13.3%), supporting the hypothesis that women have been disproportionately affected by the economic and social consequences of lockdown.

**Table 2.** Sleep loss and new occurrence during the pandemic by selected characteristics.

|  | % sleep loss during the coronavirus pandemic | % new occurrence |
| --- | --- | --- |
| **Total** | **24.7** | **20.2** |
| **Age group** | *** | *** |
| 16-24 | 29.2 | 25.0 |
| 25-44 | 31.0 | 26.3 |
| 45-64 | 26.6 | 21.3 |
| 65-74 | 16.6 | 13.7 |
| 75+ | 9.3 | 7.7 |
| **Sex** | *** | *** |
| Men | 16.5 | 13.0 |
| Women | 31.8 | 27.0 |
| **Ethnicity** | *** | * |
| British/English/Scottish/Welsh/Northern Irish (white) | 24.3 | 20.1 |
| Other white | 22.4 | 18.2 |
| Black, Asian, and minority ethnic (BAME) | 32.0 | 24.6 |
| **Highest qualification** | *** | *** |
| No qualification | 22.5 | 16.9 |
| GCSE or lower | 25.8 | 20.6 |
| A level | 26.5 | 22.3 |
| Degree | 25.7 | 21.8 |
| Unknown | 20.1 | 16.3 |
| **Live with a partner** | *** | *** |
| No | 28.8 | 22.7 |
| Yes | 22.8 | 19.2 |
| **Children in the house** | *** | *** |
| At least one child aged 0-4 | 33.6 | 28.7 |
| At least one school children aged 5-18 | 30.1 | 24.6 |
| Others | 22.4 | 18.2 |
| **Key worker** | *** | *** |
| No | 24.7 | 21.3 |
| Yes | 28.9 | 25.2 |
| Not in paid or self-employed work | 22.0 | 15.9 |
| **Has had symptoms that could be coronavirus** | *** | *** |
| No | 23.5 | 18.9 |
| Yes | 34.3 | 30.4 |
| **Feel lonely** | *** | *** |
| Hardly ever | 14.7 | 12.3 |
| Sometime | 35.9 | 30.8 |
| Often | 60.4 | 55.1 |
| **Subjective current financial situation** | *** | *** |
| Living comfortably | 15.6 | 13.5 |
| Doing alright | 23.2 | 19.9 |
| Just about getting by | 35.1 | 28.2 |
| Finding it quite difficult/very difficult | 54.8 | 48.1 |
| **Subjective future financial situation** | *** | *** |
| Better off | 22.4 | 17.9 |
| Worse off than now | 38.0 | 32.7 |
| About the same | 22.0 | 18.0 |
| **Smoker** | *** | * |
| No | 24.1 | 20.0 |
| Yes | 30.4 | 22.9 |
| **Physical activity** | *** | *** |
| No exercise | 31.2 | 23.4 |
| Had some exercise but not meet NHS guideline | 25.9 | 21.5 |
| Had exercise meet guideline | 22.6 | 18.7 |
| **Prior problem of sleep loss** | *** |  |
| No | 20.2 |  |
| Yes | 48.6 |  |
| Unknown | 25.9 |  |
| **Region** | *** | ** |
| North East | 29.3 | 22.2 |
| North West | 22.8 | 18.7 |
| Yorkshire and The Humber | 27.5 | 22.8 |
| East Midlands | 23.7 | 18.6 |
| West Midlands | 26.6 | 21.9 |
| East of England | 20.1 | 16.9 |
| London | 29.9 | 25.4 |
| South East | 24.0 | 19.2 |
| South West | 22.7 | 19.3 |
| Wales | 25.1 | 20.6 |
| Scotland | 23.7 | 18.8 |
| Northern Ireland | 26.6 | 22.6 |
| Number of respondents | 15360 | 12206 |

Source: authors' analysis, Understanding Society: COVID-19 Study, 2020.
\*\*\*p<0.001, \*\*p<0.01, \*p<0.05
Note: All proportions are weighted using sample weights. Number of respondents are unweighted.

**Figure 1:**
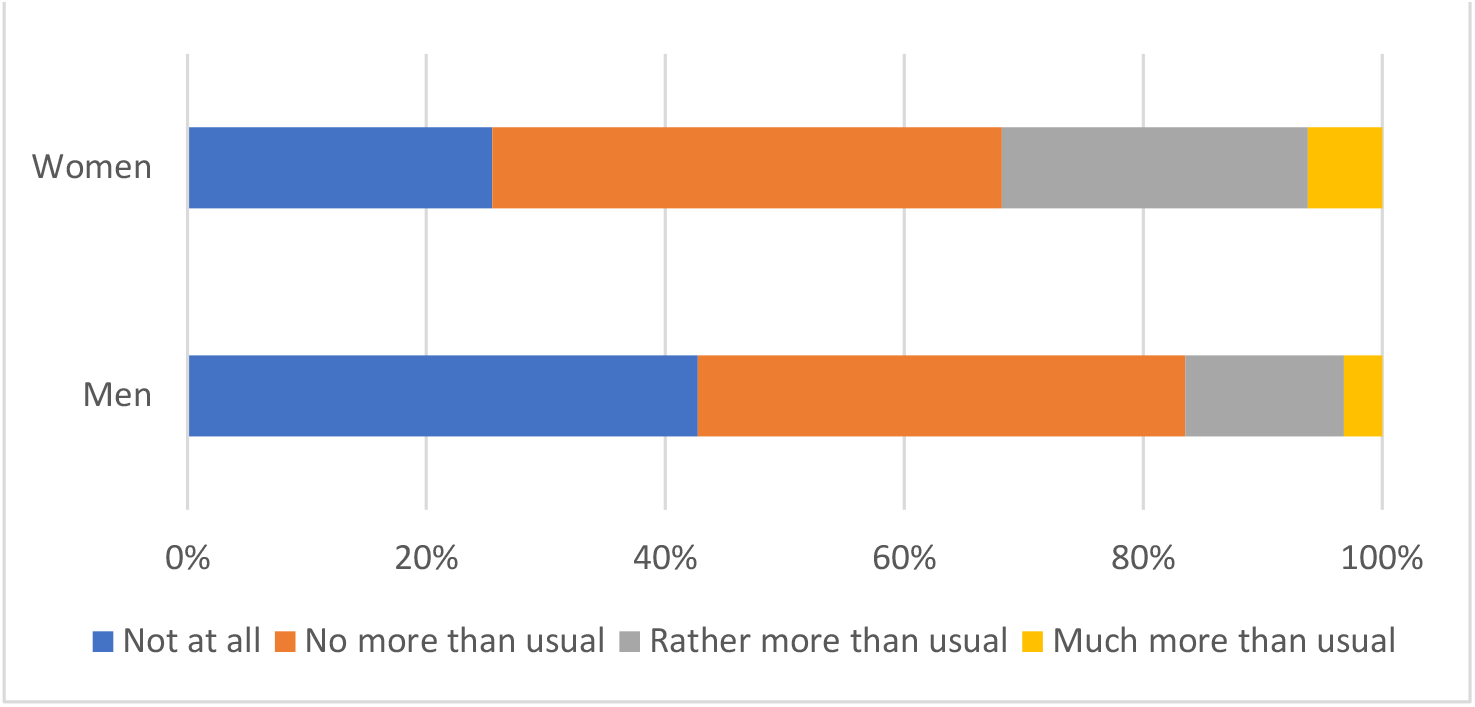
Have you recently lost much sleep over worry? By gender (%)

**Figure 2:**
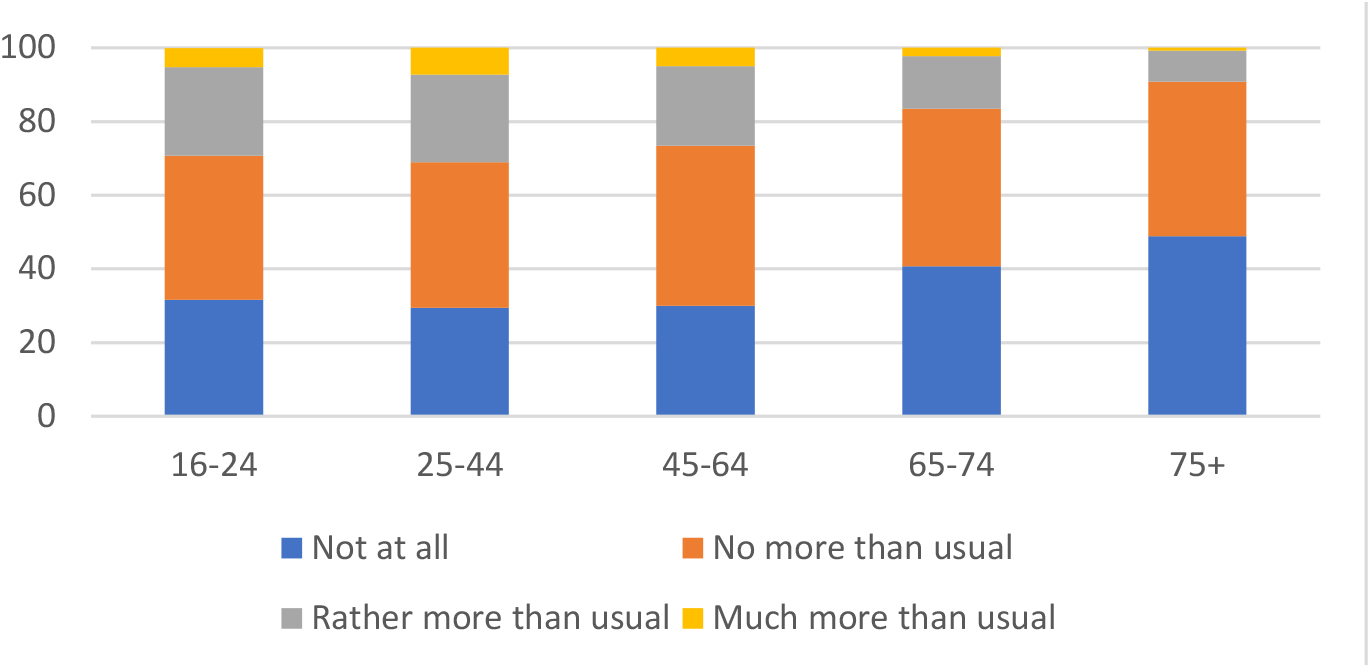
Sleep loss over worry in last few weeks by age group (%)

**Figure 3:**
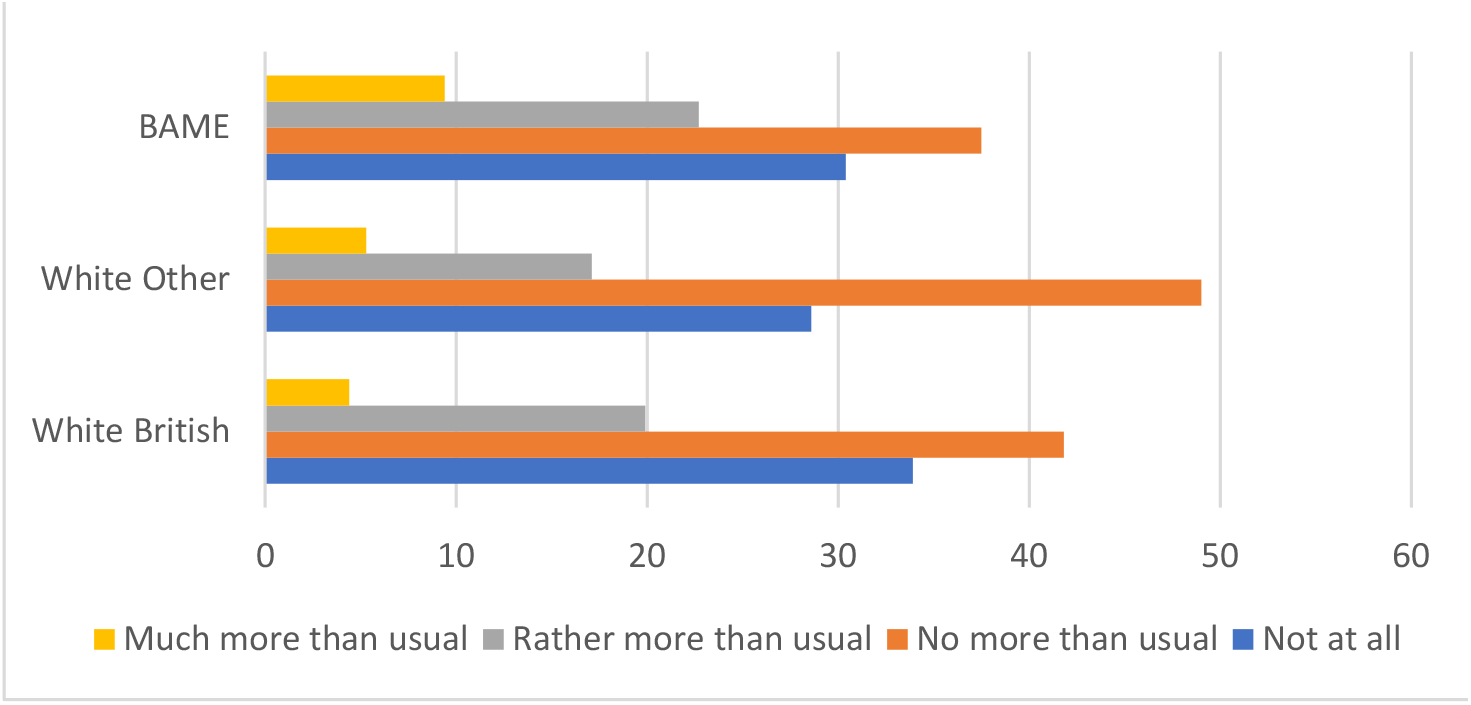
Have you recently lost much sleep over worry? By ethnic group (%)

The differences by age group further support the notion that it is not the risk of infection by Covid-19 per se that is the key factor in sleep loss, but rather the consequences associated with the policy measures taken to control the pandemic, with those aged 25-44 being more likely to report being ‘much more than usual’ to lose sleep through worry in the previous few weeks (7.3%) compared to those aged 16-24 (5.2%), 45-64 (5.0%), 65-74 (2.3%) and 75+ (0.8%). It may be that older people are more likely to lose sleep ‘normally’; however, this gradient remains even when controlling for report of sleep loss prior to the pandemic.

The results by ethnicity do, however, support the argument that the risk of infection may play a role (Figure 3); 9.4% of BAME respondents report being ‘much more than usual’ to have lost sleep through worry over the last few weeks compared to 4.4% of white respondents.

The prevalence of sleep loss has also increased compared to before the pandemic and the gaps between men and women, and individuals from different ethnic communities have widened during the epidemic (**Appendix Table A**). Amongst those clear of sleep loss before the coronavirus pandemic, one fifth of the UK population reported a new occurrence of sleep loss. The corresponding incidence rates of sleep loss among women and individuals from BAME heritage were 27.0% and 24.6% (**Table 2**). The bivariate analysis in Table 2 shows that the prevalence of sleep loss varied according to a range of factors, including those associated with Covid-19 itself, i.e. having had symptoms that could be coronavirus and being a key worker, as well as factors associated with anxiety and stress from the lockdown - including having at least one child aged 0-4 or school-aged children at home, no partner at home, being concerned about current or future financial circumstances, feeling lonely and having a prior sleep loss problem. Similar associations were also observed for the incidence rate (new occurrence of sleep loss) and the selected covariates (**Table 2**).

### Multi-variate analysis

Many of the characteristics discussed so far are likely to be interrelated; for example, those with a child aged 0-4 at home are also likely to be aged 25-44; those who are key workers are more likely to have experienced symptoms etc. In order to further unravel the picture, a series of multivariate logistic regression models were run; firstly for the population as a whole and then separately for men and women. The first two columns for Model A (**Table 3)** show the unadjusted (bi-variate) odds ratios of experiencing sleep loss by gender and ethnicity, whilst the third column shows the adjusted odds for the full model. The analysis shows that the differential impact by gender remains significant even after controlling for all other factors; the differential by ethnicity is however reversed once other factors such as being a key worker, having had symptoms, having children in the household and experiencing financial difficulties and living in London, the epicentre of the UK’s coronavirus outbreak, are taken into account. Individuals from BAME heritage are disproportionately represented in all of these groups^4^ (**Appendix Table B**).

**Table 3.** Adjusted odds ratios of sleep loss during the pandemic

|  |  | Model A<br>All respondents |  | Model B<br>Men |  | Model C<br>Women |  |
| --- | --- | --- | --- | --- | --- | --- | --- |
| **Sex** |  |  |  |  |  |  |  |
| Men (ref) |  |  |  |  |  |  |  |
| Women | 2.4*** |  | 2.0*** |  |  |  |  |
| **Ethnicity** |  |  |  |  |  |  |  |
| British/English/Scottish/Welsh/Northern Irish (white) (ref) |  |  |  |  |  |  |  |
| Other white |  | 0.9 | 0.7** | 0.8 | 0.5** | 0.9 | 0.7* |
| Black, Asian, and minority ethnic (BAME) |  | 1.4*** | 0.8** | 1.8*** | 0.9 | 1.2** | 0.7*** |
| **Age group** |  |  |  |  |  |  |  |
| 16-24 (ref) |  |  |  |  |  |  |  |
| 25-44 |  |  | 1.1 |  | 1.0 |  | 1.1 |
| 45-64 |  |  | 1.2 |  | 1.2 |  | 1.2 |
| 65-74 |  |  | 0.9 |  | 1.0 |  | 0.8 |
| 75+ |  |  | 0.6*** |  | 0.7 |  | 0.6*** |
| **Highest qualification** |  |  |  |  |  |  |  |
| No qualification (ref) |  |  |  |  |  |  |  |
| GCSE or lower |  |  | 1.0 |  | 1.0 |  | 1.0 |
| A level |  |  | 1.1 |  | 1.1 |  | 1.1 |
| Degree |  |  | 1.2* |  | 1.2 |  | 1.2 |
| Unknown |  |  | 1.0 |  | 1.0 |  | 1.0 |
| **Subjective current financial situation** |  |  |  |  |  |  |  |
| Living comfortably (ref) |  |  |  |  |  |  |  |
| Doing alright |  |  | 1.2** |  | 1.3* |  | 1.2* |
| Just about getting by |  |  | 1.7*** |  | 1.8*** |  | 1.7*** |
| Finding it quite difficult/very difficult |  |  | 3.0*** |  | 3.5*** |  | 2.7*** |
| **Subjective future financial situation** |  |  |  |  |  |  |  |
| About the same (ref) |  |  |  |  |  |  |  |
| Better off |  |  | 1.0 |  | 0.9 |  | 1.0 |
| Worse off |  |  | 1.5*** |  | 1.7*** |  | 1.3*** |
| **Live with a partner** |  |  |  |  |  |  |  |
| Yes (ref) |  |  |  |  |  |  |  |
| No |  |  | 0.8*** |  | 0.7** |  | 0.8*** |
| **Children in the house** |  |  |  |  |  |  |  |
| None (ref) |  |  |  |  |  |  |  |
| At least one child aged 0-4 |  |  | 1.3** |  | 1.5** |  | 1.1 |
| At least one school children aged 5-18 |  |  | 1.2** |  | 1.1 |  | 1.2** |
| **Key worker** |  |  |  |  |  |  |  |
| No (ref) |  |  |  |  |  |  |  |
| Yes |  |  | 1.2** |  | 1.1 |  | 1.2** |
| Not in paid or self-employed work |  |  | 1.0 |  | 0.9 |  | 1.1 |
| **Has had symptoms that could be coronavirus** |  |  |  |  |  |  |  |
| No (ref) |  |  |  |  |  |  |  |
| Yes |  |  | 1.3*** |  | 1.4*** |  | 1.2* |
| **Feel lonely** |  |  |  |  |  |  |  |
| Hardly ever (ref) |  |  |  |  |  |  |  |
| Sometime |  |  | 2.8*** |  | 3.0*** |  | 2.8*** |
| Often |  |  | 7.1*** |  | 7.3*** |  | 7.0*** |
| **Smoker** |  |  |  |  |  |  |  |
| No (ref) |  |  |  |  |  |  |  |
| Yes |  |  | 0.9* |  | 0.7* |  | 0.9 |
| **Physical activity** |  |  |  |  |  |  |  |
| No exercise (ref) |  |  |  |  |  |  |  |
| Had some exercise but not meet NHS guideline |  |  | 1.0 |  | 0.9 |  | 1.1 |
| Had exercise meet guideline |  |  | 0.9 |  | 0.8 |  | 1.1 |
| **Prior problem of sleep** |  |  |  |  |  |  |  |
| No (ref) |  |  |  |  |  |  |  |
| Yes |  |  | 2.3*** |  | 2.6*** |  | 2.1*** |
| **Region** |  |  |  |  |  |  |  |
| North East (ref) |  |  |  |  |  |  |  |
| North West |  |  | 0.9 |  | 0.7 |  | 1.1 |
| Yorkshire and The Humber |  |  | 1.1 |  | 0.7 |  | 1.3 |
| East Midlands |  |  | 0.9 |  | 0.7 |  | 0.9 |
| West Midlands |  |  | 0.9 |  | 0.9 |  | 0.9 |
| East of England |  |  | 0.9 |  | 0.8 |  | 0.9 |
| London |  |  | 1.2 |  | 1.0 |  | 1.3 |
| South East |  |  | 0.9 |  | 0.9 |  | 1.0 |
| South West |  |  | 0.9 |  | 0.8 |  | 1.0 |
| Wales |  |  | 0.9 |  | 0.7 |  | 1.0 |
| Scotland |  |  | 0.9 |  | 0.8 |  | 0.9 |
| Northern Ireland |  |  | 1.1 |  | 1.1 |  | 1.1 |
| LR Chi square | 465.30 | 32.2 | 2772.9 | 34.7 | 868.76 | 8.9 | 1487.9 |
| Chi square test | <0.001 | <0.001 | <0.001 | <0.001 | <0.001 | P<0.05 | P<0.01 |
| Pseudo R2 | 0.0271 | 0.0019 | 0.1616 | 0.0062 | 0.1545 | 0.0008 | 0.1344 |
| Number of respondents | 15360 | 15360 | 15360 | 6406 | 6406 | 8954 | 8954 |

Source: authors' analysis, Understanding Society: COVID-19 Study, 2020.
\*\*\*p<0.001, \*\*p<0.01, \*p<0.05

The analysis shows that the coronavirus infection, being a key worker, with children aged 0-4 or school-aged children at home, feeling lonely, perceived financial difficulties and worry, and being a woman, were all predictive factors of sleep loss (**Table 3**) and a new case of sleep loss (**Table 4**). The influential factors were slightly different among men (Model B) and women (model C). Among women, being a key worker was a risk factor and being a BAME was a protective factor for increased sleep loss whilst both these factors were not associated with sleep loss for men.

**Table 4.**
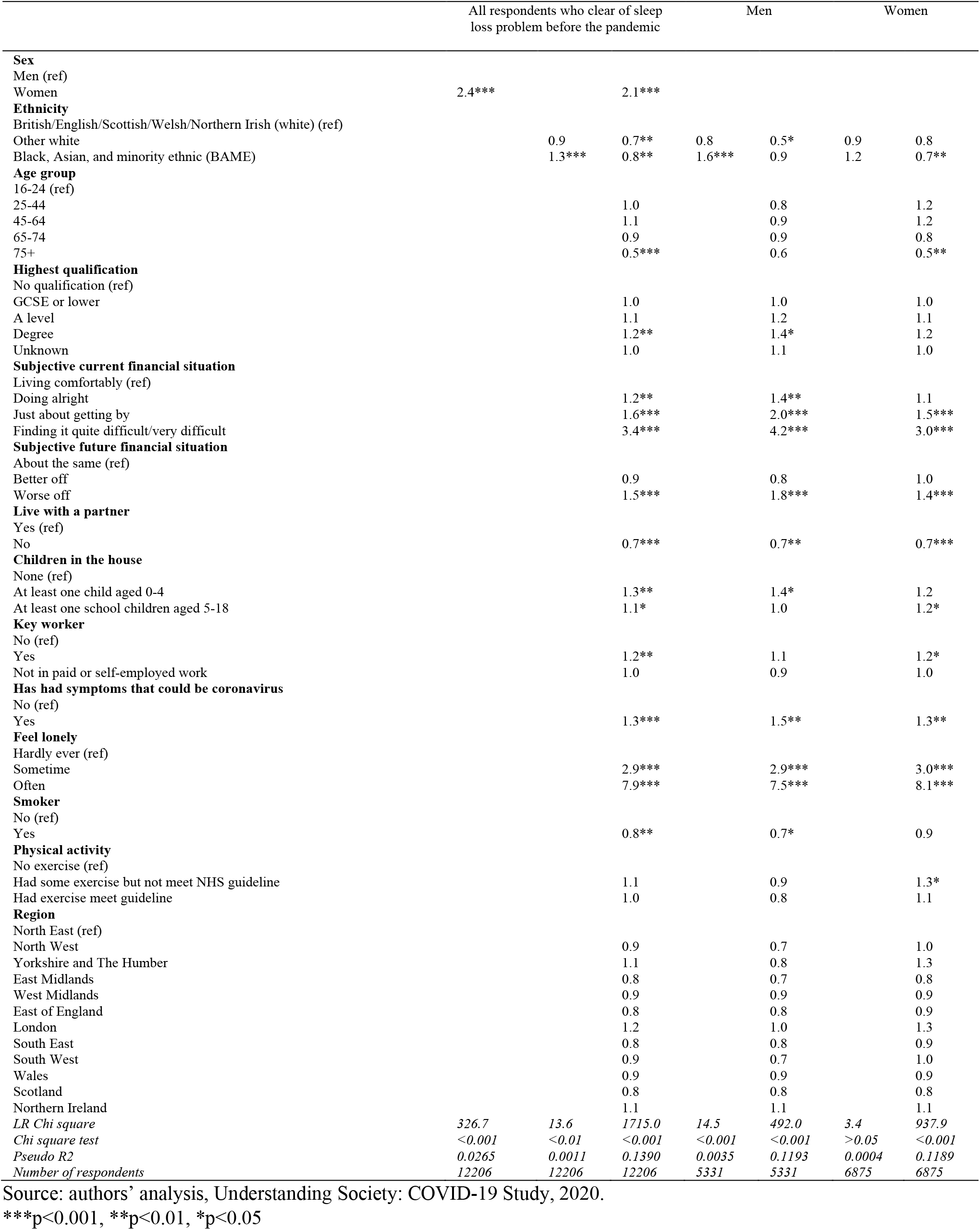
Adjusted odds ratios of new occurrence of sleep loss during the pandemic

With the move to home working and home schooling, some individuals may experience multiple stressors. Table 5 and Figures 4 & 5 present post model predictive margins (and 95% confidence intervals) for a range of exemplar respondents, illustrating the additional impact on sleep loss of cumulative stressors. Female respondents from BAME heritage with a child under 0-4 and with self-reported financial difficulties (respondent D) were nearly three times more likely to report sleep loss than on average (Respondent A) and this risk was heightened further if they were a key worker and had had symptoms (Respondent E).

**Table 5.** Post model predictive margins and 95% confidence intervals of sleep loss and new occurrence of sleep loss by respondent characteristics.

| Respondent characteristics | Predictive margin for sleep loss | Predictive margin for new occurrence of sleep loss |
| --- | --- | --- |
| Any respondent (average) | 0.21 (0.20 to 0.21) | 0.17 (0.16 to 0.17) |
| A woman | 0.26 (0.25 to 0.27) | 0.22 (0.21 to 0.23) |
| A woman, with at least one 0-4 child, with a degree and work but not a keyworker | 0.30 (0.26 to 0.33) | 0.26 (0.23 to 0.30) |
| A woman, from Black, Asian, and minority ethnic (BAME), with at least one child aged 0-4, current financial quite difficult/very difficult, future financial getting worse | 0.53 (0.47 to 0.59) | 0.52 (.044 to 0.59) |
| A woman, from Black, Asian, and minority ethnic (BAME), with at least one child aged 0-4, current financial quite difficult/very difficult, future financial getting worse, being a key worker, has had symptoms that could be coronavirus | 0.60 (0.54 to 0.67) | 0.62 (0.54 to 0.69) |

Note: predictive margins of sleep loss based on the full model in Table 3; predictive margins of new occurrence of sleep loss based on the full model in Table 4.

**Figure 4.**
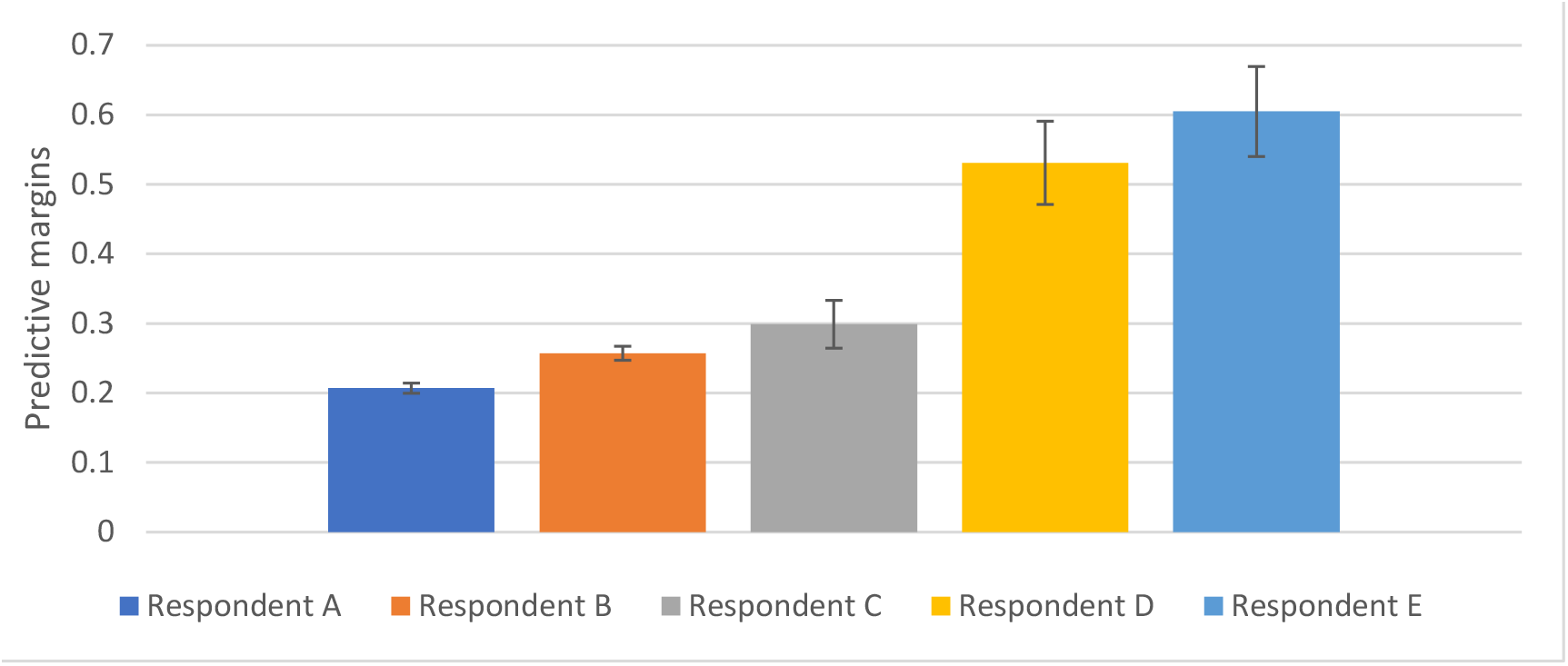
Predictive margins and 95% CI of sleep loss by respondent characteristics

**Figure 5:**
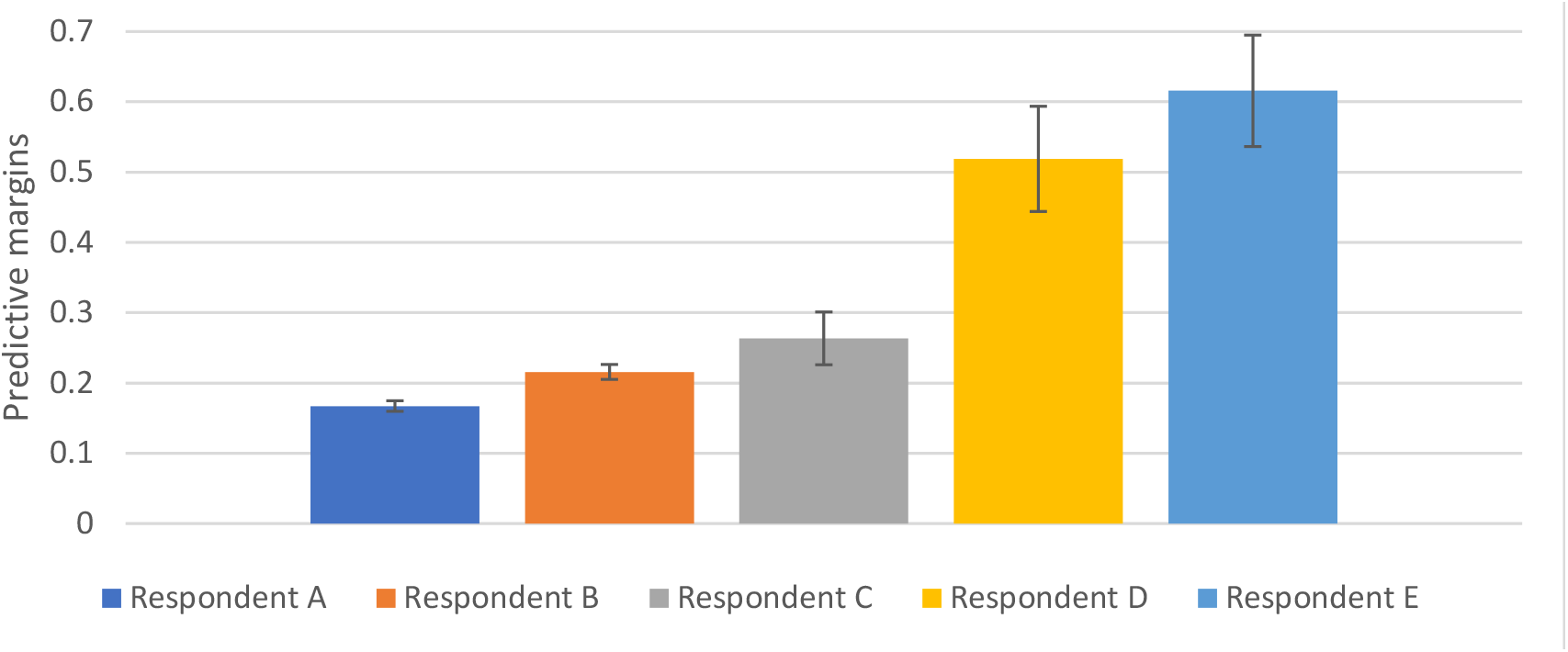
Predictive margins and 95% CI of new occurence of sleep loss by respondent characteristics Note: predictive margins of sleep loss based on the full model in Table 3; predictive margins of new occurrence of sleep loss based on the full model in Table 4. Respondent A: any respondent (average) Respondent B: a woman Respondent C: a woman, with at least one 0-4 child, with a degree and work but not a keyworker Respondent D: a woman, from Black, Asian, and minority ethnic (BAME), with at least one child aged 0-4, current financial quite difficult/very difficult, future financial getting worse Respondent E: a woman, from Black, Asian, and minority ethnic (BAME), with at least one child aged 0-4, current financial quite difficult/very difficult, future financial getting worse, being a key worker, has had symptoms that could be coronavirus

## Discussion and conclusion

This study has revealed several important findings related to sleep health during the Covid-19 pandemic. Firstly it provides robust evidence that sleep loss is affecting more people during the Covid-19 pandemic than previously, reflecting the fact that stress levels have risen due to anxieties about health, financial consequences, changes in social life and the daily routine, all of which may affect sleep homeostasis.

The study also provides evidence that women have been more vulnerable to sleep deprivation during lockdown, which is in line with previous research suggesting that women have more sleep disturbances than men (9,21), and that women are more prone to stress-related sleep disorders such as post-traumatic stress disorder and anxiety disorders (24). There is emerging evidence that mental health experiences during the Covid-19 pandemic in the UK differ between men and women, with more women suffering from anxiety in the early stages of lockdown (25). Women’s position in the labour market may increase their exposure to Covid-19, as women represent a significant majority of frontline workers in social care, education and health care (16, 26). Many parents will be affected by school closures, and requirements to balance paid work with increasing childcare and providing support to their children’s learning. However, the gendered allocation of childcare means that in many households, it is the mother who continues to provide the majority of primary care for children. Furthermore, many mid-life women find themselves juggling employment with caring responsibilities for aged parents and grandchildren (27).

People from BAME heritage had a higher prevalence and incidence rate of sleep loss than that of British White. This reflects the fact that those with BAME heritage have disproportionally higher rates of coronavirus infection, high anxiety associated with coronavirus-specific circumstances (22), are more likely to be key workers, to have dependent children, and to feel lonely. All of these are likely to increase the risk of sleep loss, with the result that once all these other factors are controlled for, being a member of BAME community was associated with a *reduced* chance of sleep loss – highlighting the complex relationship between ethnicity and sleep health.

In conclusion, the Covid-19 pandemic and the policy responses to it, including home working and schooling, have widened the disparity of sleep deprivation across gender and ethnicity, putting women and ethnic minorities at an even greater disadvantage. Disrupted and poor sleep is associated with wider mental and physical health challenges. Policy makers and health professionals need to take action now to support and promote better sleep health amongst vulnerable groups during the pandemic, averting future secondary complications.

## Data Availability

The data is available through the UK Data Service. Full reference is provided below.
University of Essex, Institute for Social and Economic Research. Understanding Society: COVID-19
Study, 2020. [data collection]. UK Data Service. 2020; SN: 8644, Available: http://doi.org/10.5255/UKDASN-8644-1.

## Author Declarations

This study used the secondary data. All relevant ethical guidelines have been followed.

---

**Appendix Table A.**
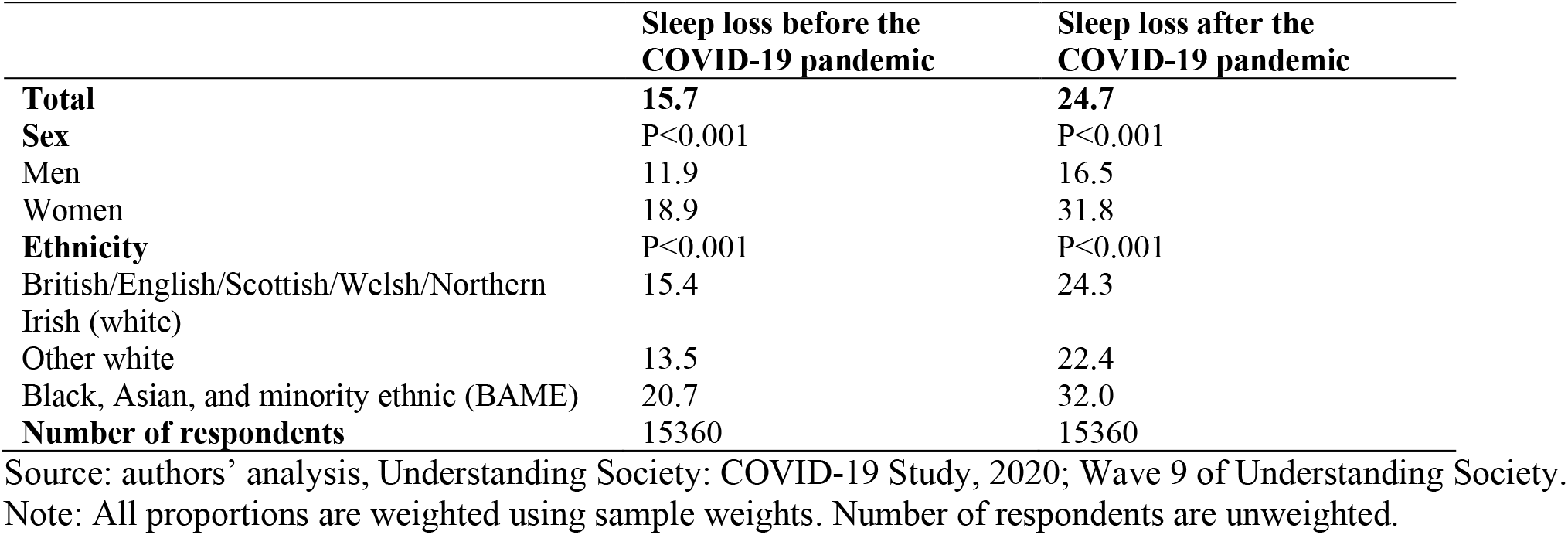
Sleep loss comparison before and after the COVID-19 pandemic by sex and ethnicity.

**Appendix Table B.** Characteristics of ethnicity.

|  | British/English/Scottish/Welsh/Northern Irish<br>(white) | Other white | Black, Asian, and minority ethnic<br>(BAME) |
| --- | --- | --- | --- |
| **Age group** | P<0.001 |  |  |
| 16-24 | 8.0 | 6.2 | 16.1 |
| 25-44 | 25.8 | 33.5 | 37.2 |
| 45-64 | 38.8 | 41.9 | 37.5 |
| 65-74 | 17.8 | 13.1 | 6.1 |
| 75+ | 9.6 | 5.3 | 3.2 |
| **Sex** | P=0.341 |  |  |
| Men | 46.2 | 42.8 | 46.8 |
| Women | 53.8 | 57.2 | 53.2 |
| **Subjective current financial situation** | P<0.01 |  |  |
| Living comfortably | 34.4 | 28.8 | 18.6 |
| Doing alright | 43.2 | 43.8 | 40.6 |
| Just about getting by | 17.1 | 18.4 | 26.3 |
| Finding it quite difficult/very difficult | 5.3 | 9.1 | 14.5 |
| **Subjective future financial situation** | P<0.01 |  |  |
| About the same | 76.7 | 65.7 | 66.8 |
| Better off | 7.5 | 10.0 | 9.2 |
| Worse off | 15.8 | 24.3 | 24.0 |
| **Children in the house** | P<0.001 |  |  |
| None | 74.4 | 64.3 | 61.0 |
| At least one child aged 0-4 | 8.0 | 6.7 | 14.3 |
| At least one school children aged 5-18 | 17.6 | 29.0 | 24.7 |
| **Key worker** | P<0.01 |  |  |
| No | 33.5 | 43.0 | 30.1 |
| Yes | 26.7 | 24.6 | 31.1 |
| Not in paid or self-employed work | 39.8 | 32.4 | 38.8 |
| **Has had symptoms that could be coronavirus** | P<0.01 |  |  |
| No | 88.7 | 86.5 | 83.4 |
| Yes | 11.3 | 13.5 | 16.6 |
| **Feel lonely** | P<0.01 |  |  |
| Hardly ever | 63.0 | 59.4 | 54.4 |
| Sometime | 28.8 | 29.5 | 35.9 |
| Often | 8.2 | 11.1 | 9.7 |
| **Prior sleep loss** | P<0.001 |  |  |
| No | 83.7 | 85.2 | 74.4 |
| Yes | 15.4 | 13.5 | 20.7 |
| **Region** | P<0.001 |  |  |
| North East (ref) | 4.5 | 0.9 | 0.8 |
| North West | 11.4 | 5.5 | 11.0 |
| Yorkshire and The Humber | 8.8 | 3.8 | 7.7 |
| East Midlands | 8.2 | 5.5 | 5.4 |
| West Midlands | 9.1 | 3.5 | 9.6 |
| East of England | 10.5 | 10.0 | 8.0 |
| London | 7.4 | 24.1 | 38.1 |
| South East | 15.1 | 15.0 | 9.7 |
| South West | 10.0 | 6.9 | 4.6 |
| Wales | 4.8 | 4.9 | 1.1 |
| Scotland | 8.4 | 4.6 | 3.7 |
| Northern Ireland | 1.8 | 15.3 | 0.3 |

Source: authors' analysis, Understanding Society: COVID-19 Study, 2020.
Note: All proportions are weighted using sample weights.

## Footnotes

1 Current UK guidelines for physical activity (21) recommend that adults should spend at least 150 minutes per week in moderately intense physical activity, in bouts of ten minutes or longer, or 75 minutes per week of vigorous physical exercise, or a combination of the two.

2 According to Department of Health and Social Care guidance on testing eligibility (2020) (20), key workers are people whose jobs are vital to public health and safety during the coronavirus lockdown. The list includes health and social care, e.g. all NHS staff, frontline health and social care staff such as doctors, nurses, plus support and specialist staff in the health and social care sector; education and childcare, including social workers; food and other necessary goods; key public services; local and national government; utility workers; public safety and national security; and transport.

3 In the first few weeks of the pandemic in the UK, the rate of infection was much higher in London than elsewhere in the country.

4 A range of interaction effects with ethnicity were investigated but none were significant and thus are not included in the final model.

